# Temporal dynamics in viral shedding and transmissibility of COVID-19

**DOI:** 10.1101/2020.03.15.20036707

**Authors:** Xi He, Eric HY Lau, Peng Wu, Xilong Deng, Jian Wang, Xinxin Hao, Yiu Chung Lau, Jessica Y Wong, Yujuan Guan, Xinghua Tan, Xiaoneng Mo, Yanqing Chen, Baolin Liao, Weilie Chen, Fengyu Hu, Qing Zhang, Mingqiu Zhong, Yanrong Wu, Lingzhai Zhao, Fuchun Zhang, Benjamin J Cowling, Fang Li, Gabriel M Leung

**Author notes:** Corresponding author: Eric HY Lau. joint first authors who contributed equally (Xi He, Eric HY Lau). joint senior authors who contributed equally (Benjamin J Cowling, Fang Li, Gabriel M Leung).

## Abstract

We report temporal patterns of viral shedding in 94 laboratory-confirmed COVID-19 patients and modelled COVID-19 infectiousness profile from a separate sample of 77 infector-infectee transmission pairs. We observed the highest viral load in throat swabs at the time of symptom onset, and inferred that infectiousness peaked on or before symptom onset. We estimated that 44% of transmission could occur before first symptoms of the index. Disease control measures should be adjusted to account for probable substantial pre-symptomatic transmission.

## MAIN TEXT

The disease now called COVID-19 was first identified in December 2019 in a cluster of cases of viral pneumonia linked to a wet market in Wuhan.^1^ SARS-CoV-2 infection spreads efficiently between people with a basic reproductive number in the range 2.0 to 2.5 in Wuhan.^2-4^ The modes of transmission have not been determined, but infection is presumed to spread mainly through respiratory droplets and fomites, similar to other respiratory viruses. The World Health Organization declared COVID-19 a global pandemic on 11 March 2020.

The effectiveness of control measures depends on several key epidemiological parameters (Figure 1a). The serial interval is defined as the duration between symptom onsets of successive cases in a transmission chain. The incubation period is defined as the time between infection and onset of symptoms. The incubation period can differ from individual to individual, and the serial intervals can vary between transmission chains, and this variation can be summarized by the incubation period distribution and the serial interval distribution respectively. If the observed mean serial interval is shorter than the observed mean incubation period, this indicates that a significant portion of transmission may have occurred before infected persons have developed symptoms. A large proportion of pre-symptomatic transmission would likely reduce the effectiveness of control or preventive measures that are directly or indirectly initiated by symptom onset, such as isolation, contact tracing for quarantine, and enhanced hygiene or use of face masks for symptomatic persons. Failing to consider pre-symptomatic transmission would likely overestimate the effect of such measures when devising control strategies.

**Figure 1.**
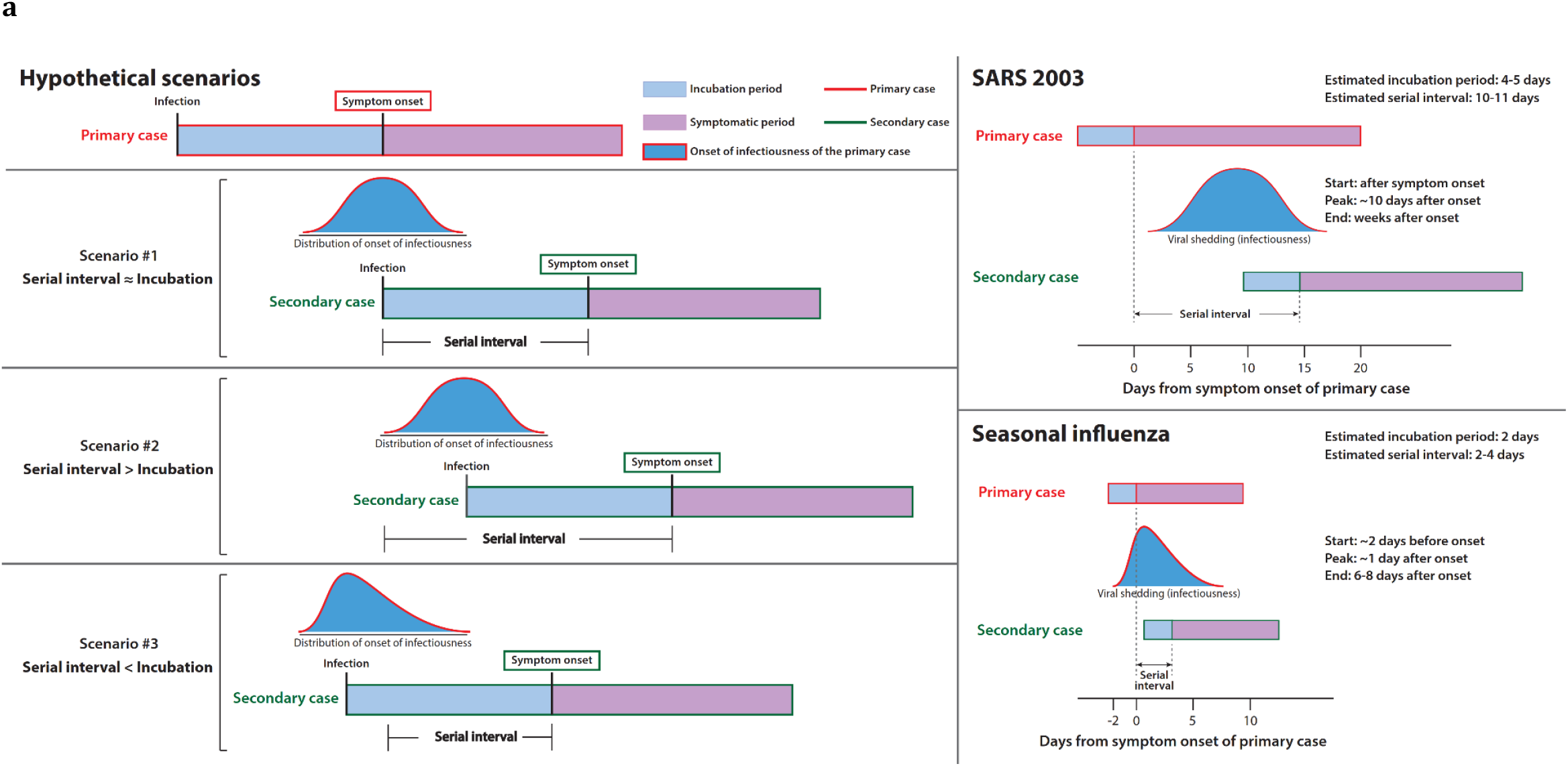
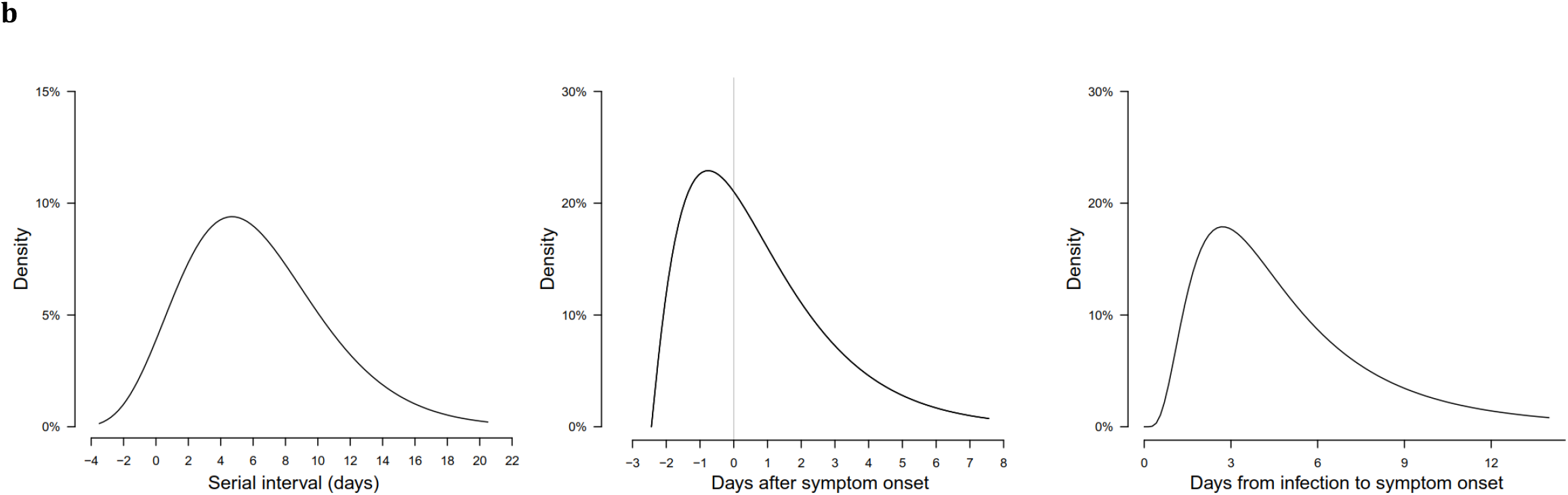
**a**, A schematic of the relation between different time periods in the transmission of infectious disease. **b**, Estimated serial interval distribution (left), inferred infectiousness profile (middle) and assumed incubation period (right).

As a comparison, the Severe Acute Respiratory Syndrome (SARS) coronavirus was notable because infectiousness increased around 7-10 days after illness onset.^5,6^ Once containment measures were effectively implemented, including isolation of cases and quarantine of their close contacts, it was possible to reduce substantially onwards transmission of infection (Figure 1a).^7,8^ In contrast, seasonal influenza is characterized by increase in infectiousness shortly around or even before illness onset.^9^ For SARS-CoV-2, there remains very limited information to date on the dynamics of contagiousness over time.

In this study, we compared clinical data on virus shedding with separate epidemiologic data on incubation periods and serial intervals between cases in transmission chains, to draw inferences on infectiousness profiles.

Among 94 laboratory-confirmed COVID-19 patients admitted to Guangzhou Eighth People’s Hospital, 47/94 (50%) were male and the median age was 47 years. 61/93 (66%) of the patients were moderately ill (with fever and/or respiratory symptoms and radiographic evidence of pneumonia) but none were classified as “severe” or “critical” on hospital admission, although 20/94 (21%) of the patients deteriorated to a severe or critical condition during hospitalization.

A total of 414 throat swabs were collected from these 94 patients (median = 4 swabs per patient), from the day of illness onset up to 32 days after onset. We detected high viral loads soon after illness onset, which gradually decreased towards the detection limit at about 21 days after onset (Figure 2). There was no obvious difference in viral loads across sex, age groups and disease severity.

**Figure 2.**
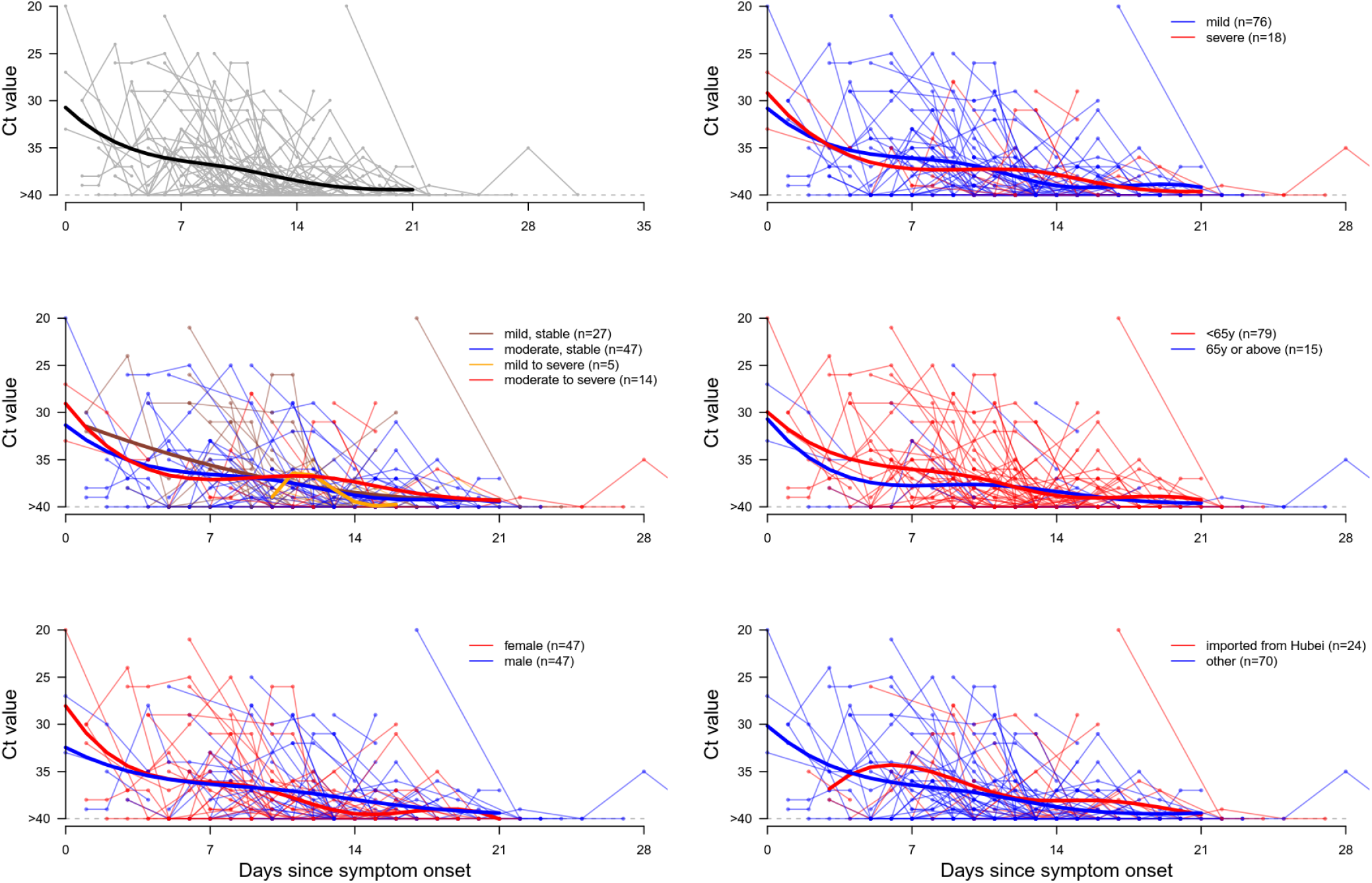
Viral load (Ct values) detected by RT-PCR in throat swabs from patients infected with SARS-CoV-2 (N=94), overall and stratified by disease severity, sex, age group and link to Hubei province. The detection limit was Ct = 40 which was used to indicate negative samples. The black line showed the trend in viral load using smoothing spline.

Separately, based on 77 transmission pairs obtained from publicly available sources within and outside mainland China, the serial interval was estimated to have a mean of 5.8 days (95% confidence interval [CI] = 4.8 to 6.8) and median of 5.2 days (95% CI = 4.1 to 6.4) based on a fitted gamma distribution (Figure 1b). Assuming an incubation period with a mean of 5.2 days from a separate study of early COVID-19 cases,^2^ we inferred that infectiousness started from 2.5 days before symptom onset and reached its peak at 0.6 days before symptom onset (Figure 1b). The proportion of transmission before symptom onset (area under the curve) was 44%. Infectiousness was estimated to decline relatively quickly within 7 days of illness onset. Viral load data was not used in the estimation but showed similar monotonic decreasing pattern after symptom onset.

On sensitivity analysis, instead of estimating the start of infectiousness from the model, we fixed the start of infectiousness at 4 and 7 days before symptom onset respectively. We estimated that peak infectiousness was reached at about 0-2 days before symptom onset, and the proportion of transmission occurring before illness onset was 52% and 46% respectively.

Finally, simulation showed that when infectiousness started at the same time as symptom onset, there would be a small proportion of short serial intervals (e.g. < 2 days). The proportion would become larger if infectiousness were to start before symptom onset, where the resulting serial intervals would resemble more those observed (Figure 1b).

Here we used detailed information on the timing of symptom onsets in transmission pairs to infer the infectiousness profile of COVID-19. We found substantial transmission potential for patients who have not yet shown symptoms, with 44% of transmission prior to symptom onset. This should however be interpreted in the context of most cases being isolated after symptom onset, preventing some post-symptomatic transmission that might otherwise have occurred. Nevertheless it is clear that pre-symptomatic transmission is most likely occurring at significant frequencies.

Our modeling analysis suggests that viral shedding may begin 2-3 days before showing first symptoms. After symptom onset, viral loads decreased monotonically, consistent with a recent study of 17 patients from another hospital in Guangdong.^10^ We could not identify patient subgroups which demonstrated a different viral shedding pattern and our results is likely applicable to COVID-19 patients generally. Whereas a recent study from Wuhan reported that virus was detected for a median of 20 days (up to 37 days amongst survivors) after symptom onset,^11^ infectiousness may decline significantly after 10 days.^12^ Together this supports our findings that the infectiousness profile may more closely resemble that of influenza than of SARS (Figure 1a), although we did not have data on viral shedding before symptom onset for COVID-19.^9,13^ Our results are also supported by reports on asymptomatic and pre-symptomatic transmission.^14,15^

For a reproductive number of 2.5,^2^ contact tracing and isolation alone are less likely to be successful if more than 30% of transmission occurred before symptom onset, unless >90% of the contacts can be traced.^16^ Such high ascertainment is more likely achievable if the definition of contacts covered 2-3 days prior to symptom onset of the index case, as is being done in Hong Kong since 22 February and in mainland China since 21 February. Even when control strategy is shifting away from containment to mitigation as the pandemic unfolds, contact tracing would still be an important measure, such as when there are super-spreading events that may occur in frail and high-risk groups such as nursing homes or hospitals. Preliminary indications of pre-symptomatic infectivity of COVID-19 cases has already prompted enhanced pre-emptive preventive measures in the nosocomial setting and in the community, especially for high risk groups such as older adults and those with pre-existing medical conditions. With a substantial proportion of pre-symptomatic transmission, measures such as enhanced personal hygiene among the general population and social distancing would likely be the key instruments for disease control in the community.

Our study has several limitations. First, the infectiousness profile was inferred from infector-infectee pairs and could be affected by the isolation of some cases, as well as behavioral changes after symptom onset such as enhanced personal hygiene and self-isolation. Second, we reviewed available information on known serial intervals, but the observations we identified may have been biased towards shorter intervals because reporting of more recent exposure to sick persons before symptom onsets is more prominent. This may have shifted the inferred infectiousness profile towards negative (i.e. transmission before symptom onset). Nevertheless in our dataset of transmission pairs we were able to capture relatively long serial intervals (e.g. > 10 days), hence the impact on the inferred infectiousness was likely limited. Our simulation also shows how infectiousness starting before symptom onset would be more consistent with the observed serial interval distribution.

In conclusion, we found that viral shedding of laboratory-confirmed COVID-19 patients peaked on or before symptom onset, and a substantial proportion of transmission likely occurred before first symptoms in the index case. More inclusive criteria for contact tracing to capture potential transmission events 2-3 days before symptom onset should be urgently considered for effective control of the outbreak.

## Methods

### Sources of Data

Guangzhou Eighth People’s Hospital in Guangdong, China was designated as one of the specialized hospitals for treating COVID-19 patients at both city and provincial levels on 20 Jan 2020. Since then many COVID-19 cases were admitted via fever clinics, the hospital emergency room or after confirmation of cases from community epidemiological investigation carried out by the Guangzhou Center for Disease Control and Prevention, or transferred from other hospitals. The first confirmed COVID-19 case was admitted on 21 Jan 2020, but in the initial phase suspected cases have also been admitted. We identified all suspected and confirmed COVID-19 cases admitted from 21 Jan 2020 to 14 Feb 2020, and collected throat swabs from them. Patients included those who travelled from Wuhan or Hubei to Guangzhou, and local cases, ranging from asymptomatic, mild to severe cases on admission.

The samples were tested by N-gene-specific quantitative reverse-transcriptase– polymerase-chain-reaction (RT-PCR) assay as previously described.^17^ To understand the temporal dynamics of viral shedding and exclude non-confirmed COVID-19 cases, we selected 94 patients who had at least one positive results (Cycle threshold [Ct] value < 40) in their throat samples. Serial samples were collected from some but not all patients for clinical monitoring purposes.

We collected information reported on possible human-to-human transmission pairs of laboratory-confirmed COVID-19 patients from publicly available sources including announcements made by government health agencies and media reports in mainland China and countries/regions outside of China. A transmission pair was defined as two confirmed COVID-19 cases who were identified in the epidemiologic investigation showing a clear epidemiologic link with each other and one case (infectee) was highly likely infected by the other (infector), by fulfilling the following criteria: 1) the infectee did not report a travel history to an area affected by COVID-19 or any contact with other confirmed or suspected COVID-19 cases except for the infector within 14 days before symptom onset; and 2) the infector and infectee were not identified in a patient cluster where other COVID-19 cases had also been confirmed; and 3) the infector and infectee pair did not share a common source of exposure to a COVID-19 case or a place where there were COVID-19 case(s) reported. We excluded possible transmission pairs without a clear exposure history reported prior to symptom onset. Data of possible transmission pairs of COVID-19 were extracted, including age, sex, location, date of symptom onset, type or relationship between the pair cases, and time of contact of the cases.

### Statistical analysis

We analyzed two separate data sets – clinical and epidemiologic – to assess pre-symptomatic infectiousness. First, we assessed longitudinal viral shedding data from laboratory-confirmed COVID-19 patients starting from symptom onsets, where viral shedding during the first few days after illness onset can be compared with the inferred infectiousness. Second, the serial intervals from clear transmission chains, combined with information on the incubation period distribution, were used to infer the infectiousness profile as described below.

We present SARS-CoV-2 viral loads in the throat swabs of each patient by day of symptom onset. A smoothing spline was fitted to the Ct values to summarize the overall trend. We also compared the viral load by disease severity, age, sex and travel history from Hubei. We fitted a gamma distribution to the transmission pairs data to estimate the serial interval distribution.

We used a published estimate of the incubation period distribution to infer infectiousness with respect to symptom onset, from the first 425 COVID-19 patients in Wuhan with detailed exposure history.^2^ We considered that infected cases would become infectious at a certain time point before or after illness onset (t_S1_). Infectiousness, i.e. transmission probability to a secondary case, would then increase until reaching its peak (Figure 1). The transmission event would occur at time t_I_ with a probability described by the infectiousness profile β(t_I_ - t_S1_) relative to the illness onset date, assuming a gamma distribution. The secondary case would then show symptoms at time t_S2_, after the incubation period that is assumed to follow a lognormal distribution g(t_S2_ - t_I_). Hence the observed serial intervals distribution f(t_S2_ - t_S1_) would be the convolution between the infectiousness profile and incubation period distribution. We constructed a likelihood function based on the convolution which was fitted to the observed serial intervals, allowing for the start of infectiousness around symptom onset. Parameters were estimated using maximum likelihood. We also performed sensitivity analyses by fixing the start of infectiousness at day 4 and 7 respectively before symptom onset and inferred the infectiousness profile.

As an additional check, we simulated the expected serial intervals assuming the same aforementioned incubation period but two different infectiousness profiles, where infectiousness started on the same day and from 2 days before symptom onset respectively. A recent study isolated live infectious SARS-CoV-2 virus from COVID-19 patients up to 10 days after symptom onset,^12^ thus we assumed the same duration of infectiousness. We also assumed that infectiousness peaked on the day of symptom onset. The timing of transmission to secondary cases were simulated according to the infectiousness profile using a lognormal and exponential distribution respectively, where the serial intervals were estimated as the sum of the onset to transmission interval and the incubation period. We present the distribution of the serial intervals over 10,000 simulations.

All statistical analyses were conducted in R version 3.6.2 (R Development Core Team, Vienna, Austria).

## Ethics approval

Data collection and analysis were required by the National Health Commission of the People’s Republic of China to be part of a continuing public health outbreak investigation.

## Data Availability

Detailed transmission pairs data in this study are given in the supplementary information
and viral shedding data will be available upon request and approval by a data access
committee. The data access committee comprises leadership of the Guangzhou Eighth
People’s Hospital and the Guangzhou Health Commission; there is no restriction to data
access.

## Acknowledgements

This work was supported by Department of Science and Technology of Guangdong Province (Project No #2020B111108001) and a commissioned grant from the Health and Medical Research Fund from the Government of the Hong Kong Special Administrative Region.

## Contributions

XH, EHYL, PW, BJC, FL and GML conceived and designed the study. XH, XD, JW, YG, XT, XM, YC and BL were responsible for clinical care and collected all biomaterials. WC and FH carried out laboratory testing. QZ, MZ and YW collected and collated linked clinical-epidemiologic data. LZ, FZ and FL supervised and coordinated all aspects of the study at Guangzhou Eighth People’s Hospital. PW, XH, YCL and JYW collected and verified all infector-infectee transmission data. EHYL, BJC and GML wrote the first draft. All authors contributed to data interpretation, critical revision of the manuscript and approved the final version of the manuscript.

